# Gut microbial changes associated with obesity in youth with type 1 diabetes

**DOI:** 10.1101/2023.12.01.23299251

**Authors:** Heba M Ismail, Dimuthu Perera, Rabindra Mandal, Linda A DiMeglio, Carmella Evans-Molina, Tamara Hannon, Joseph Petrosino, Sarah Javornick CreGreen, Nathan W Schmidt

## Abstract

Obesity is increasingly prevalent in type 1 diabetes (T1D) and is associated with management problems and higher risk for diabetes complications. Gut microbiome changes have been described separately in each of T1D and obesity, however, it is unknown to what extent gut microbiome changes are seen when obesity and T1D concomitantly occur. Objective: To describe the gut microbiome and microbial metabolite changes associated with obesity in T1D. We hypothesized significant gut microbial and metabolite differences between T1D youth who are lean (BMI: 5-<85%) vs. those with obesity (BMI: ≥95%). Methods: We analyzed stool samples for gut microbial (using metagenomic shotgun sequencing) and short-chain fatty acid (SCFA) metabolite differences in lean (n=27) and obese (n=21) T1D youth. The mean±SD age was 15.3±2.2yrs, A1c 7.8±1.3%, diabetes duration 5.1±4.4yrs, 42.0% females, and 94.0% were White. Linear discriminant analysis (LDA) effect size (LEfSe) was used to identify taxa that best discriminated between the BMI groups. Results: Bacterial community composition showed differences in species type (β-diversity) by BMI group (p=0.013). At the genus level, there was a higher ratio of *Prevotella* to *Bacteroides* in the obese group (p=0.0058). LEfSe analysis showed a differential distribution of significantly abundant taxa in either the lean or obese groups, including increased relative abundance of *Prevotella copri*, among other taxa in the obese group. Functional profiling showed that pathways associated with decreased insulin sensitivity were upregulated in the obese group. Stool SCFAs (acetate, propionate and butyrate) were higher in the obese compared to the lean group (p<0.05 for all). Conclusions: Our findings identify gut microbiome, microbial metabolite and functional pathways differences associated with obesity in T1D. These findings could be helpful in identifying gut microbiome targeted therapies to manage obesity in T1D.

## Introduction

Obesity is prevalent in children and adolescents with type 1 diabetes (T1D) and can be as high as 46.1% in some populations [1, 2]. Epidemiological studies have also shown increasing prevalence of obesity among adults with T1D [3, 4]. This is concerning because obesity is associated with reduced insulin sensitivity [5, 6], higher exogenous insulin needs, chronic inflammation and higher risk for hypoglycemia, dyslipidemia and long-term complications [2, 7]. Therefore, it is important to identify potential contributing factors to obesity in T1D.

T1D development is thought to be influenced by the intestinal microbiome [8]. Children with T1D, and islet antibody positive relatives who later progress, exhibit gut dysbiosis (microbial imbalance) and lower gram-positive to gram-negative gut bacterial ratios, compared to healthy children [9-18]. In addition, the gut microbiome may influence T1D progression through the production of microbial metabolites. Indeed, it has been shown that the gut microbiome of children diagnosed with T1D contains fewer fermenters that produce butyrate, a short chain fatty acid (SCFA) with anti-inflammatory actions that also enhances the immune regulatory response [10-22].

Separately, gut microbiome differences and dysbioses have also been associated with overweight, obesity and insulin resistance in children and adults without diabetes [23-27]. These microbiome changes appear to be distinct from the changes seen in individuals with T1D. For example, animal and human studies suggest higher relative abundance of specific taxa (such as *Actinobacteria and Bacteroidetes*) associated with and linked to changes in secondary bile acid (BA) and steroid acid biosynthesis in obese adolescents and adults [25], while T1D studies show lower counts of *Firmicutes and Bifidobacteria* [8, 10]. Others have also proposed that a BA-gut microbiome axis contributes to insulin sensitivity and obesity [28].

Therefore, there is substantial evidence that the gut microbiome differs in healthy individuals compared to individuals with T1D, and in lean compared to obese individuals without T1D. However, the gut microbiomes of children with T1D and obesity compared to those with T1D who are lean have not been described. Here, we hypothesized that the gut microbiome composition and metabolite profile is different in lean and obese youth with T1D.

## Methods

After institutional review board approval was obtained (Institutional Review Board approved protocol number 1908640459), we enrolled obese and lean youth with T1D. Prior to any procedures, participants provided written assent and parents provided written informed consent. All methods were performed in accordance with the relevant guidelines and regulations. Participants were identified from our clinics and approached for their interest. The study was designed as a pilot study to determine effect sizes for future studies with an enrollment goal of 20 in each BMI group. We enrolled a total of 48 individuals with T1D (27 lean and 21 with obesity).

A lean BMI was defined as between the 5th-85th% for age and sex and obese as a BMI of ≥95th% for age and sex. These groups were chosen to allow for a clearer comparison of the two of BMI percentile categories at both ends of the BMI spectrum. Other inclusion criteria included age between 11 and 18 years at the time of enrollment. Exclusion criteria included: known monogenic forms of diabetes or Type 2 diabetes (confirmed clinically and by genetic/antibody testing); history of ongoing infection or antibiotic treatment within the past 3 months; history of immune-compromise, recurrent infections, steroid intake (inhaled or oral forms) or other immunosuppressant use in the past 6 months; history of chronic gastrointestinal disease, possible or confirmed celiac disease; participation in any research intervention trials within the past 3 months; and history of treatment or use of metformin, a type 2 diabetes medication that affects the gut microbiome [28].

Stool sample kits (consisting of gloves, Zymo feces catcher, RNA/DNA shield fecal collection tubes with preservative [for DNA sequencing] and without [for SCFA analysis] and freezer packs) were shipped to participants, who were asked to collect a single stool sample at home within 1-3 days prior to their study visit. Once delivered at the study visit, stool samples were immediately stored at -80°C at the research facility until the time of analysis. Clinical data pertaining to glycemic control and diabetes duration were obtained from participant records.

### DNA extraction and sequencing

We characterized the gut microbiome in participant stool samples using whole genome shotgun (WGS) sequencing. DNA was extracted using the Qiagen MagAttract PowerSoil DNA KF kit (Formerly MOBio PowerSoil DNA Kit) using a KingFisher robot. DNA quality was evaluated visually via gel electrophoresis and quantified using a Qubit 3.0 fluorometer (Thermo- Fischer, Waltham, MA, USA). Libraries were prepared using an Illumina Nextera library preparation kit with an in-house protocol (Illumina, San Diego, CA, USA). Paired-end sequencing (150 bp x 2) was done on a NextSeq 500 in medium-output mode at Microbiome Insights.

Demultiplexed raw fastq sequences were first processed using BBDuk [29] (version 38.82) to quality trim, remove Illumina adapters, and filter PhiX reads (standard Illumina spike in). Trimming parameters were set to a k-mer length of 19 and a minimum Phred quality score of 25. Reads with a minimum average Phred quality score <23 and length < 50 bp after trimming were discarded. Trimmed FASTQs were mapped to a combined PhiX and human reference genome (GCF_000001405.26) database using BBMap [29] (version 38.82) to determine and remove human/PhiX reads. Taxonomy assignments were determined using MetaPhlAn3, which generates bacterial taxon abundances/sample using clade-specific marker genes. MetaPhlAn3 has been shown to be more accurate than other pipelines with fewer false positive and false negative results [30]. Functional gene (KO, EC, UniRef 90) and metabolic pathway (metacyc) profiles were generated using HUMAnN3 [30]. The MetaPhlAn3 [31, 32] recommended workflow with a modification to the read mapping tool was used for taxonomic profiling. Processed FASTQ reads were mapped against the MetaPhlAn3 marker gene database (mpa_v30_CHOCOPhlAn_201901) using BBMap before processing through the MetaPhlAn3 algorithm. The default relative abundance and estimated counts table per kingdom per sample were further processed using a combination of MetaPhlAn3 utility scripts and in-house code designed to construct tables with improved readability for downstream statistical analysis. Functional gene profiling was done using HUMAnN3 [31, 32], following the standard recommended workflow with modifications to the nucleotide and translated alignment steps. Briefly, nucleotide alignment was performed using BBMap, generating a HUMAnN3-compatible SAM file. The translated alignment step was performed using DIAMOND [32] (version 0.9.26). The default pathway abundance table as well as gene family abundance outputs per sample were then processed using a combination of HUMAnN3 utility scripts and in-house code designed to construct tables with improved readability. Using the legacy KEGG databases included with HUMAnN 1.0, genefamilies outputs were also converted to KEGG Pathways. This was done by processing the genefamilies default output tables through the ‘humann’ script specifying the ‘pathways-database’ for the KEGG Pathways. All outputs were reported in RPK (Reads Per Kilobase) units and normalized to relative abundance units. Outputs were split and stratified by taxa and unstratified metagenome tables.

We assessed measures of α-diversity using both richness (number of species and Shannon Diversity Index) and evenness (Simpson index) measures. Differences in the β- diversity (a measure of species diversity) was compared using PERMANOVA and Bray-Curtis dissimilarities.

### Stool microbial metabolite measurements

Given the significant tole of SCFAs in health and disease, we opted to measure SCFA levels in stool. SCFA extraction and data were analyzed using gas chromatography-flame ionization detection (GC-FID), [33]. Stool material were resuspended in MilliQ-grade H2O, and homogenized using MP Bio FastPrep, for 1 minute at 4.0 m/s. 5M HCl was added to acidify fecal suspensions to a final pH of 2.0. Acidified fecal suspensions were incubated and centrifuged at 10,000 RPM to separate the supernatant. Fecal supernatants were spiked with 2- Ethylbutyric acid for a final concentration of 1 mM. Extracted SCFA supernatants were stored in 2-ml GC vials, with glass inserts. SCFAs were detected using gas chromatography (Thermo Trace 1310), coupled to a flame ionization detector (Thermo). Our SCFA column used was ‘Thermo TG-WAXMS A GC Column, 30 m, 0.32 mm, 0.25 um’ [33]. Individual fatty acids were plotted, and concentrations were normalized to the amount of input material (mmol SCFA/ kg Cecal Fecal).

### Dietary Recall

A 24-hour dietary recall was obtained using the Automated Self-Administered 24-Hour (ASA24) dietary assessment tool [34, 35], a free validated web-based tool for administering automatically coded, self-administered recalls/food records. Parents/caregivers were asked to complete this for the 24-hour time period prior to stool collection. The online survey also provides analytic output files for different macro and micro-nutrients. We analyzed data collected on the main dietary components (carbohydrate, fat and protein intake) as well as fiber intake and total caloric intake per day. Here, we assessed differences in dietary intake to determine if differences could possibly contribute to differences in the gut microbiome composition or function.

### Statistical and Bioinformatics Analysis

Baseline demographics data were compared between the two BMI groups. Student’s T-Test was used for comparison of continuous variables and Kruskal-Wallis was used for comparison of categorical data. P-value of <0.05 was used for statistical significance level.

Downstream analysis of microbiome trends as they relate to sample metadata was performed using ATIMA [https://atima.research.bcm.edu], a stand-alone tool for analyzing and visualizing microbiome data sets. The software is a web application combining publicly available R packages [36-38] with purpose-written code to import sample data and identify trends in taxa abundance, alpha diversity, and beta diversity. Categorical variables were evaluated using the non-parametric Mann-Whitney U [39] and Kruskal-Wallis tests [40] for variables with two or three and more groups, respectively. Relationships with continuous variables were tested using R’s base function for linear regression models (‘lm’). Differences in between sample (beta) diversity were assessed by calculating Bray-Curtis dissimilarity matrices and visualizing using PCoA ordination. The ADONIS function in the R package VEGAN (version 2.5.5) was used to perform pairwise comparisons and estimate PERMANOVA p-values of the community composition between BMI groups [41, 42]. All p-values are adjusted for multiple comparisons with Benjamini and Hochberg’s formula to control for the false discovery rate.

For differential abundance analysis of taxonomic and pathway profiles between the BMI groups, raw input data was entered into MaAsLin2 (**M**icrobiome **M**ultivariable **A**ssociation with **L**inear Models) [43]. Covariates of interest and that were used for adjustments included sex, age, HbA1c and diabetes duration. Linear discriminant analysis (LDA) effect size (LEfSe) was used to identify the taxa that best discriminated between the lean and obese groups, using the nonparametric Kruskal-Wallis sum-rank test to detect significantly different taxa or pathways between groups. LDA significance threshold was set at >±2. T-tests were used to compare the SCFA levels between the BMI groups and Kruskal-Wallis to compare certain bacterial ratios.

## Results

We enrolled 48 participants with T1D (27 lean; 21 obese). The overall mean (±SD) age was 15.3±2.2 years, diabetes duration 5.1±4.4 years, HbA1c 7.8±1.3%, and 42.0% were female, 94.0% were Non-Hispanic Whites. The average daily insulin dose was 38.4±28.0 and only 11 individuals were on insulin pumps while the remaining were on multiple daily injections. Demographic data by BMI group are included in Supplemental Table 1.

### Bacterial Community Analysis

There were no significant differences in measures of α-diversity by BMI group, Figure 1A. There was a significant difference in β-diversity observed between lean and obese BMI groups in PCoA plots with Bray-Curtis dissimilarity metrics (p = 0.013; Figure 1B). This appeared to be driven mostly by *Provetellaceae* and *Bacteroidaceae*. After adjusting for baseline demographics, diabetes duration and HbA1c, this difference disappeared (p=0.319).

**Figure 1:**
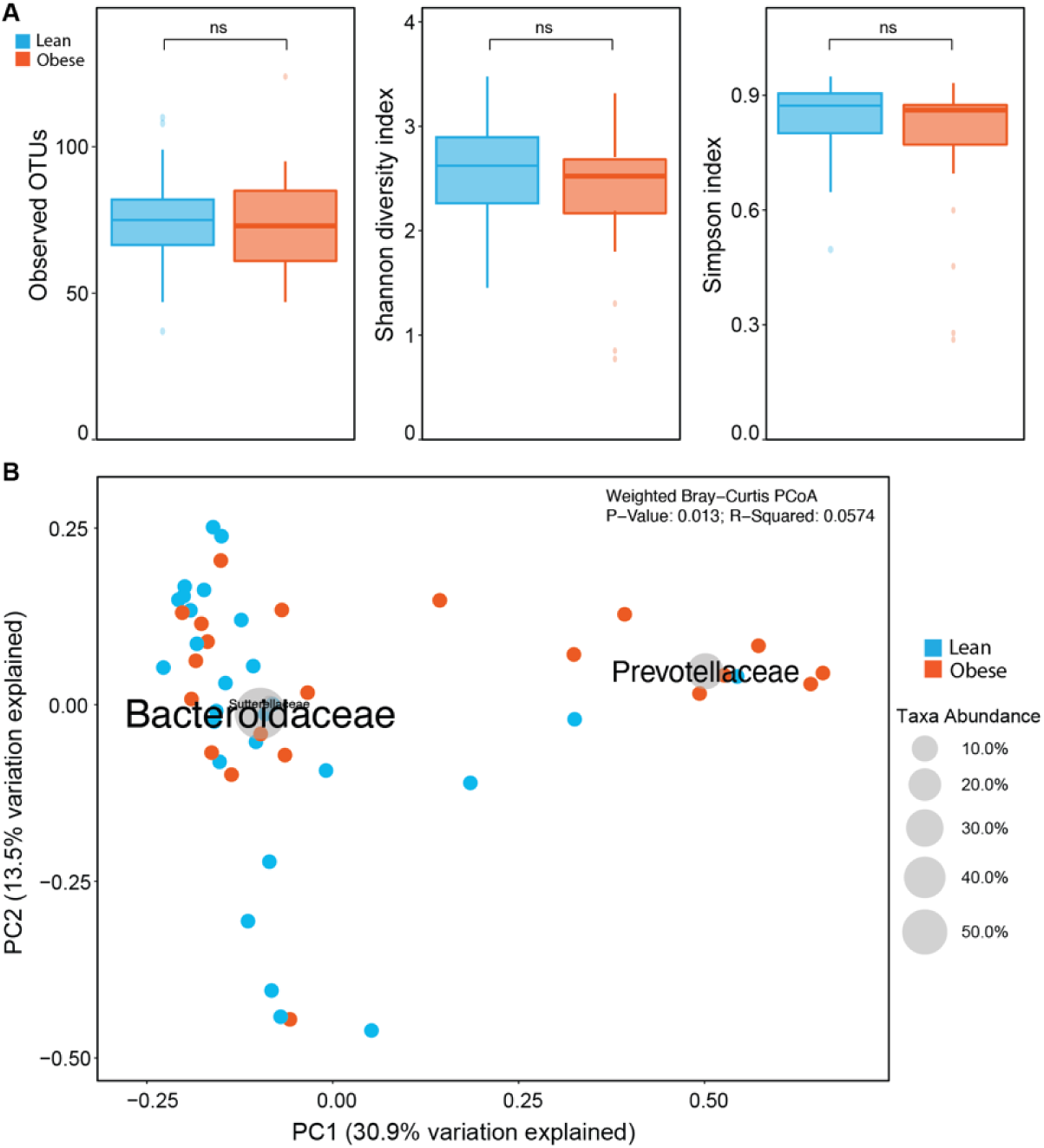
Measures of alpha- and beta-diversity in youth with type 1 diabetes. A) Observed OTUs, Shannon diversity and Simpson index were not significantly different between lean and obese BMI group samples. B) Bray-Curtis PCoA ordination of stool samples from lean and obese BMI group showed a significant difference between the groups (p-value = 0.013). Overlaid bacterial taxonomy bi-plot shows the differences driven by *Prevotellaceae* and *Bacteroidaceae*.

At the genus level, the relative abundance of *Bacteroides* was higher in the lean group, while *Prevotella* was relatively more abundant in the obese group (p<0.05 before adjustments), Supplemental Figure 1. This significant difference disappeared after adjustment for baseline variables. Comparison of the *Firmicutes* to *Bacteroidetes* ratio (F/B), a ratio commonly lower in those with or at risk for T1D did not yield a significant difference, Figure 2A. However, we also assessed the *Prevotella* to *Bacteroides* ratio (P/B), which has been studied in obesity, and found a significantly higher P/B in the obese T1D group (p=0.0058), Figure 2B.

**Figure 2:**
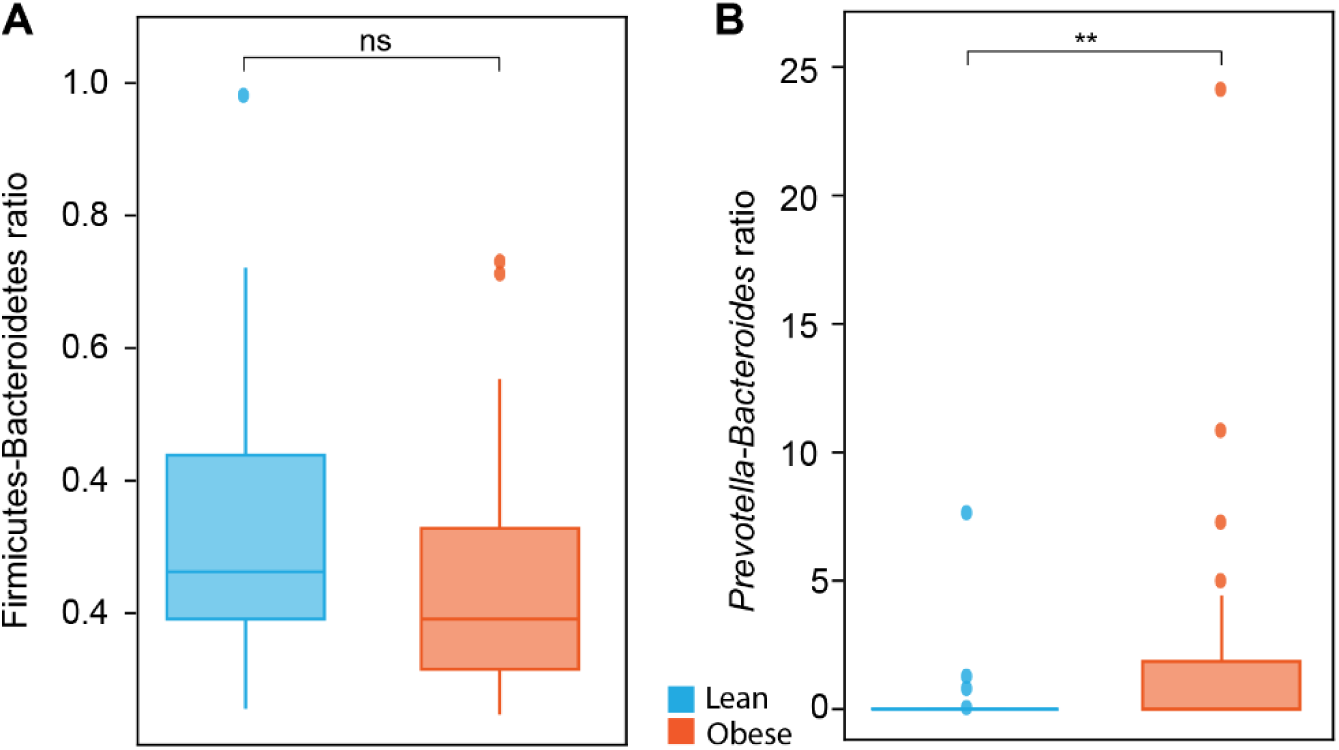
Comparison of the ratio between taxa commonly associated with T1D and obesity studies. A) Firmicutes to Bacteroidetes ratio (F/B) – no significant difference between BMI groups. B) *Prevotella* to *Bacteroides* ratio (P/B) – significantly higher in the obese T1D group (p=0.0058).

Deeper analysis using LEfSe revealed significant differences in the relative abundance of several taxa. For increased stringency, we filtered the taxa to only retain those that met an LDA threshold of ±2, and the log (10) transformed score is shown to demonstrate the effect size (Figure. 3A). Notable was an increased relative abundance of several fermenters in the obese group (*Prevotella copri, Bacteroides stercoris, Parabacteroides merdae, Holdemenalla biformis* and *Bifidobacterium angulatum)* while the lean group showed an increased relative abundance of several SCFA producers (*Anaerotruncus colihominis, Clostridium sp. CAG:167, Bifidobacterium pseudocatenulatum, Eubacterium sp. CAG:251, Eubacterium eligens, Bacteroides plebius, B. thetaiotaomicron and B. ovatus).* Differences remained after adjustment for baseline variables. In a post-hoc analysis of a subset of the individuals matched for age, sex and race (16 in each BMI group), we observed repeat shared differences in specific species with the larger group of 48 samples, Figure 3B.

**Figure 3:**
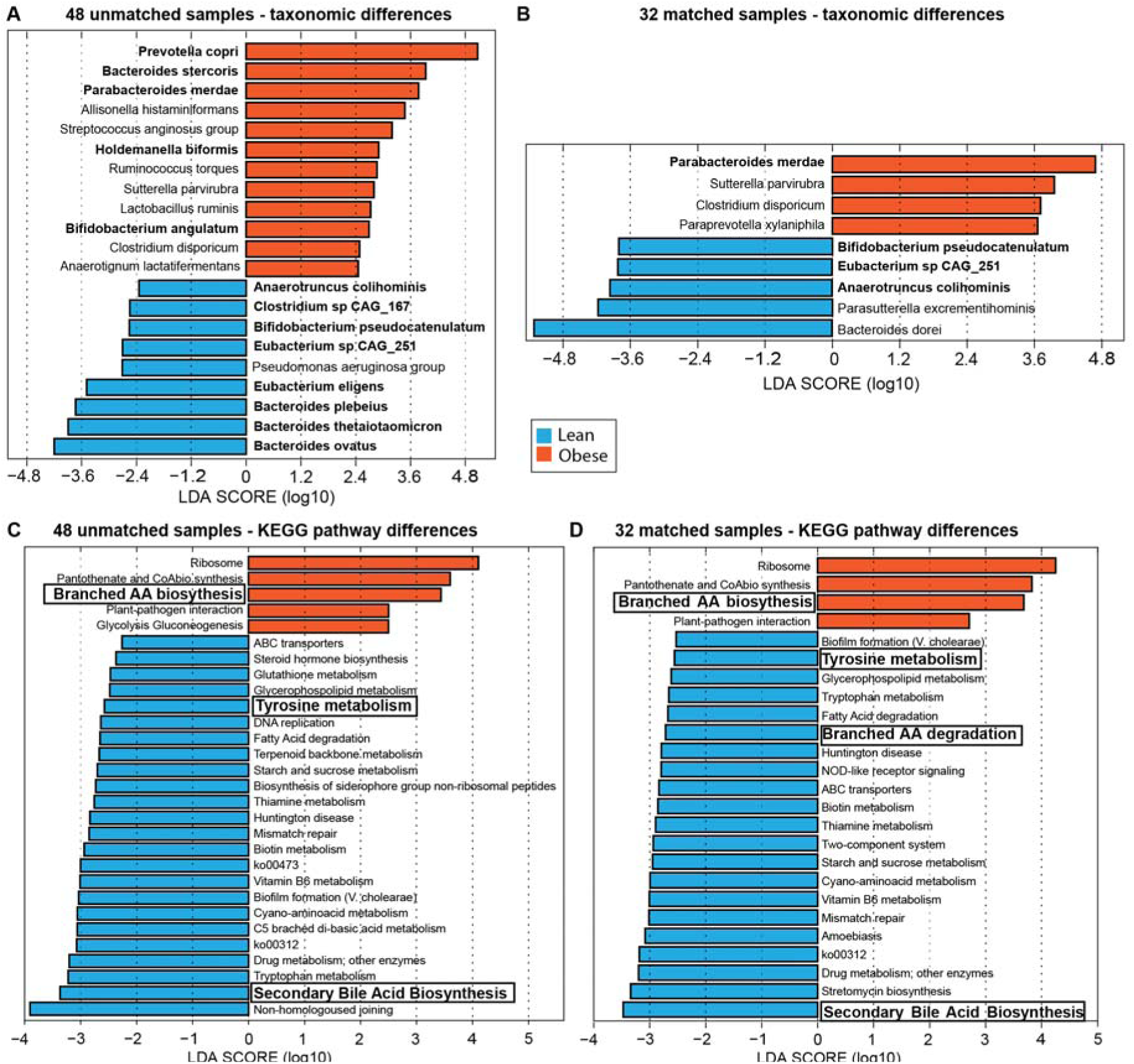
Linear discriminant analysis effect size of differentially abundant bacterial species and KEGG pathways in youth with type 1 diabetes. Horizontal bars represent the effect size for each pathway: orange enrichment in obese group, and blue enrichment in lean group. LDA score cutoff of 2 was used. **A-B)** Differential abundance of bacterial species in total 48 sample set (A) and matched 32 sample set (B). No per-sample normalization done. **C-D)** Differential abundance of KEGG pathways in total 48 sample set (C) and matched 32 sample set (D). Per-sample normalization used and Unmapped or Unintegrated pathways removed.

### Functional pathways differences

When assessing functional pathways, several findings were observed, whether in the unmatched 48 samples (Figures 3C) or the 32 matched samples (Figure 3D). Notable differences were an upregulation of branched amino acid (BCAA) synthesis pathways in the obese group (associated with insulin resistance) vs upregulation in the lean group of BCAA degradation, tyrosine metabolism (higher levels of which have been associated with insulin resistance in those with fatty liver), increased secondary BA biosynthesis (associated with improved insulin sensitivity and secretion), and increased metabolism of several Vitamin Bs. Adjustments for covariates did not change the results. There were no fermentation pathway differences seen.

### Microbial metabolite differences

We compared normalized stool SCFA levels in the lean and obese groups. The obese BMI group showed significantly higher levels of the 3 main SCFAs, acetate, butyrate and propionate, Figure 4A. We next assessed differences in dietary intake to determine if this had possibly contributed to differences in the gut microbiome composition or function. There were no significant differences in the main dietary component intake for carbohydrates, saturated fats or proteins and no differences in added sugars, absolute fiber intake and fiber intake normalized per 1000Kcal (Figure 4B).

**Figure 4:**
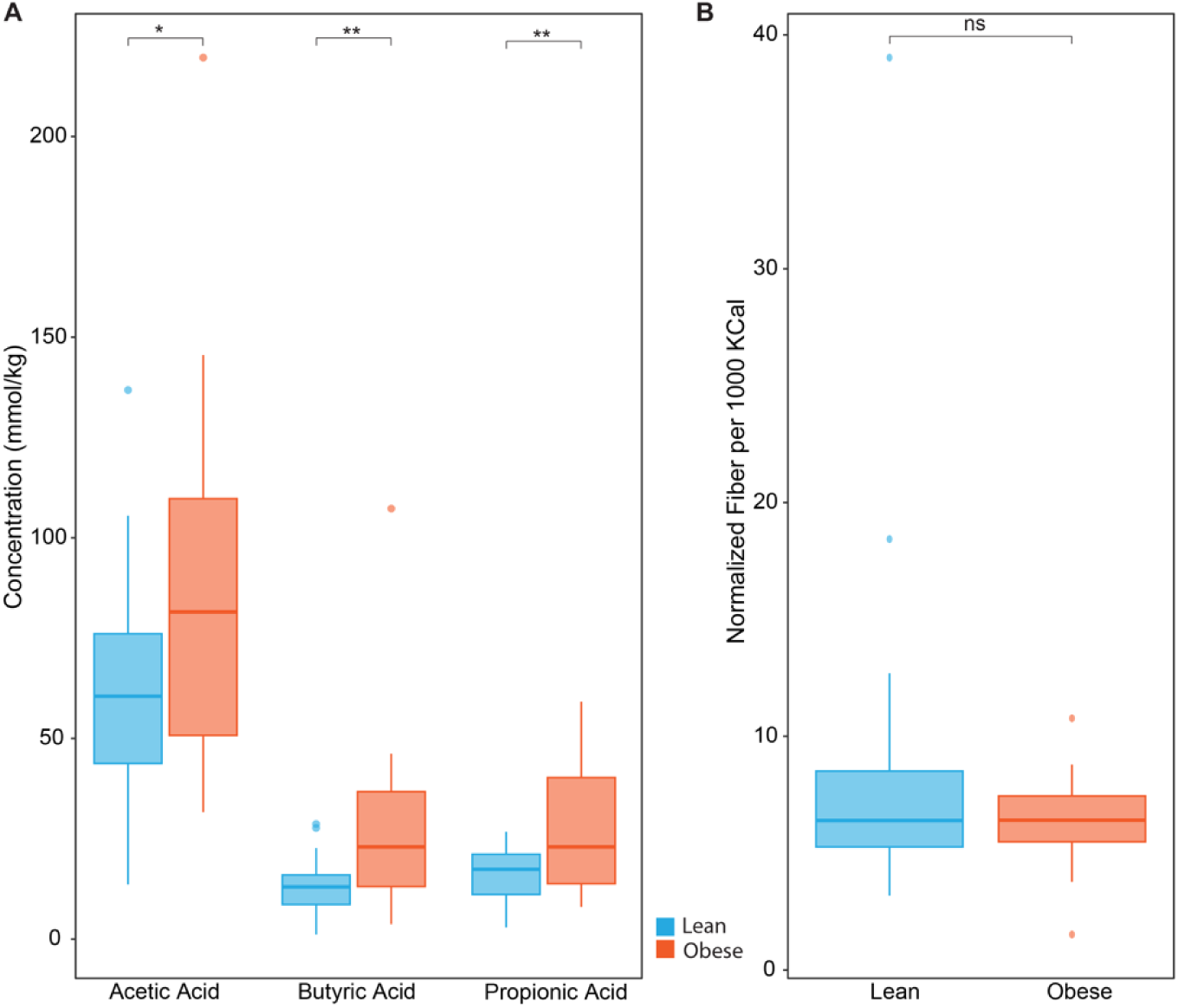
Differences in SCFA levels and fiber intake in youth with type 1 diabetes. A) Differences in levels of the 3 main SCFAs, acetate, butyrate and propionate, between lean and obese BMI groups. **B)** Comparison of fiber intake normalized per 1000Kcal between lean and obese BMI groups.

## Discussion

In this study we examined the differences in the gut microbiome and microbial metabolite composition in youth with T1D who are lean and those with T1D and obesity. This is the first study to examine these differences. Our results show youth with T1D and obesity have significant differences in the differential abundance of several taxa, significant differences in the functional pathways as well as differences in fecal SCFA levels compared with youth with T1D who are lean. Most notable were the increased relative abundance of several taxa including *Prevotella* (*P.) copri*, upregulation of BCAA biosynthesis pathways as well as higher fecal levels of all three main SCFAs in the obese group. In the lean group, there was a higher relative abundance of the *Bacteroides* species, as well as upregulation of several pathways associated with improved insulin sensitivity such as the secondary BA biosynthesis pathway and tyrosine metabolism. In a subgroup of individuals matched for age, sex and race, there was further upregulation of BCAA degradation in the lean group.

The most interesting was the increased relative abundance of *P. copri* as well as the significantly increased P/B ratio in the obese group, which is an agreement with what other studies have shown [44]. In a study by Dong et al [45], the researchers showed that a high P/B ratio was an independent risk factor for obesity and that this was associated with changes in the brain’s reward centers regulating food intake. In a study of adult Chinese individuals [46], the researchers also described increased relative abundance of *Prevotella* overall in the obese group along with an increased mean relative frequency of *P. copri* in the obese group. Further, other studies have shown that *P. copri* can perpetuate an inflammatory environment [47], and chronic systemic inflammation is a common feature of obesity and insulin resistance in general. We also assessed the F/B ratio, a common ratio studied in either type 1 diabetes or obesity. We observed no difference in the F/B ratio. Indeed, while preclinical studies have demonstrated increased *Firmicutes* and decreased *Bacteroidetes* in the microbiome of obese mice relative to that of lean mice [48, 49], human studies have demonstrated either a very small association or no associations at all [50-53].

We saw differences in several functional pathways between the lean and obese groups, and these differences persisted after adjustments for baseline covariates. Further, differences persisted in the subgroup of 32 individuals matched for age, sex and race. We found that BCAA metabolism and degradation pathways were differentially enriched in the microbiome of lean individuals vs an upregulation of biosynthesis in the gut microbiome of those with obesity. Indeed, it has been shown that *Prevotella*-rich microbiomes are associated with insulin resistance through the production of BCAAs [54]. Several studies have shown that higher levels of BCAAs are associated with obesity and insulin resistance [55], including studies of children and adolescents [56]. Further, in a nested case-control analysis of 2422 individuals followed for 12 years in the Framingham Offspring Study, plasma concentrations of five branched-chain and aromatic amino acids (isoleucine, leucine, valine, tyrosine, and phenylalanine) were identified as predictors of future development of diabetes, independent of traditional risk factors [57].

We observed upregulation of pathways associated with secondary BA biosynthesis in the lean group with T1D, which is consistent with what has been published in the literature. Higher secondary BA levels have been shown to improve insulin sensitivity by blocking the farsenoid X receptor [28]. Animal and human studies suggest higher relative abundance of taxa associated with and linked to changes in secondary BA biosynthesis in lean adolescents and adults [46, 58]. Others have also proposed that a BA-gut microbiome axis contributes to insulin sensitivity and obesity [59, 60].

Tyrosine levels have been shown to be positively associated with measures of insulin resistance in adults with non-alcoholic fatty liver disease [61] and interestingly, we saw increased tyrosine metabolism in the gut microbiome of lean individuals with T1D. While this does not necessarily translate into lower circulating tyrosine levels, it is potentially suggestive of improved insulin sensitivity in the lean group. Vitamin B synthetic pathways were also upregulated in the gut microbiome of lean youth with T1D, but not in that of those with obesity and T1D. Indeed, severe obesity has been associated with an absolute deficiency in bacterial biotin producers and transporters, whose abundances correlate with host metabolic and inflammatory phenotypes. Further, others have found that gut microbiota in the lean gut seems to be more involved with vitamin B6 biosynthesis and offer more vitamin B6 for absorption [62].

Finally, we did not observe a difference in pathways associated with fermentation. This is possibly related to the abundance of fermenters in the microbiome of both BMI groups, suggesting that these pathways may be equally upregulated in both groups. However, we found that fecal SCFA levels were higher in the obese compared to the lean groups. This was an interesting and intriguing finding at first as we had expected the SCFA levels to be higher in the lean group given the beneficial effects of SCFAs. However, it has been shown that rodent models of genetic obesity and overweight adult human studies have shown higher stool SCFA levels in the obese groups compared to the lean groups, and that this was potentially linked to increased dietary harvest of energy by the gut microbiota [63-65]. Further, Muller et al. [66] showed that circulating, rather than fecal, SCFAs were associated with GLP-1 concentrations, whole-body lipolysis, and peripheral insulin sensitivity in humans, suggesting that circulating levels are more directly linked to metabolic health. It is also possible that the metabolic and functional pathways are different in children vs. adults and in obese T1D youth vs. lean T1D youth. Of further relevance is a study by de la Cuesta et al [67], who reported higher fecal SCFAs in the obese group and found this to be positively correlated with *P. copri* in those obese individuals. They further suggested that this may be related to poor or lower SCFA efficiency absorption. Chen et al [68] described that dominance of *Prevotella* versus *Bacteroides* in fecal innocula differentially impacted *in vitro* fermentation profiles of SCFAs from fibers with different chemical structures, resulting in high production of total SCFAs (with propionate as the major product) and significantly promoted one single *Prevotella* OTU.

Strengths of this study include the use of sophisticated measures such as WGS sequencing which enables taxonomic analysis down to the bacterial species level, and functional analysis of the microbiome and the potential effects on different metabolic pathways via gene content analysis. Further, we used very stringent methods in our analysis of the data. We also used MaAsLin2, which is catered for sparse and compositional microbiome data analysis and uses a variety of models to fit the data (LM, NEGBIN, ZINB, CPLM) [43].

This study has limitations. We often found the diet recall data collected in this study to be incomplete and could have been possibly skewed by recall bias which may have contributed to the lack of differences seen in main dietary components between the lean and obese groups, which is a nature of diet recall studies. This study describes an association and does not necessarily indicate causation. However, other studies have shown that certain microbial metabolites and linked pathways are activated and potentially contribute to obesity. Future directions should include assessing for measures of insulin resistance. However, a larger study, based on results from this study, is currently underway to validate our findings and further includes an intervention to alter the gut microbiome of individuals with obesity and T1D (NCT05414409).

In summary, our study is the first to examine and show differences in the relative abundance of several bacterial taxa as well as differences in several functional pathways associated with either a lean BMI or obesity in youth with T1D. Further, we found significantly higher SCFA levels in the stool of individuals with obesity and T1D. These findings may provide insights into the pathophysiology of disease. Further evidence will be needed to verify the role of the changed gut microbiota in the development of obesity in patients with T1D.

## Data Availability

The data can be made available on request from the corresponding author, HMI.

## Contribution statement

HMI conceptualized the study. HMI, DP, RM, SJC and NWS analyzed and interpreted the data, and wrote the manuscript. LAD, CEM, TH, JP contributed to the design, interpreted the data and reviewed/edited the manuscript.

## Funding/Acknowledgements

This study received support from the National Institutes of Health, National Center for Advancing Translational Sciences, Clinical and Translational Sciences Award, Grant Numbers, KL2TR002530 (A Carroll, PI), and UL1TR002529 (A. Shekhar, PI). We also acknowledge support from the Board of Directors of the Indiana University Health Values Fund for Research Award and the Indiana Clinical and Translational Sciences Institute funded, in part by Grant U54TR002529 from the National Institutes of Health, National Center for Advancing Translational Sciences, Clinical and Translational Sciences Award; the Indiana Clinical and Translational Sciences Institute funded, in part by Award Number ULITR002529 from the National Institutes of Health, National Center for Advancing Translational Sciences, Clinical and Translational Sciences Award; the Pilot and Feasibility Grant from the Indiana Center for Diabetes and Metabolic Diseases (P30DK097512); the National Institute Of Diabetes And Digestive And Kidney Diseases of the National Institutes of Health under Award Number K23DK129799, the Doris Duke Charitable Foundation through the COVID- 19 Fund to Retain Clinical Scientists Collaborative Grant Program (Grant 2021258) and The John Templeton Foundation (Grant 62288). The content is solely the responsibility of the authors and does not necessarily represent the official views of the National Institutes of Health or other funding agencies.

