## Supplementary material for "Gut microbial changes associated with obesity in youth with type 1 diabetes": Suppl table and figure

**Supplemental Materials:**

**Supplemental Table 1: Participant demographics.**

|  | Lean  (n=27) | Obese  (n=21) |
| --- | --- | --- |
| Mean±SD Age (yrs) | 15.0±2.2 | 15.6±2.2 |
| Mean±SD BMI (%) | 56.5±21.7 | 97.9±1.3 |
| Mean±SD Diabetes Duration (yrs) | 4.8±4.4 | 5.6±4.6 |
| Sex (% Females) | 48 | 33 |
| %Non-Hispanic Whites | 97 | 95 |
| Mean±SD A1c (%) | 7.6±1.4 | 8.0±1.3 |

**Supplemental Figure 1: Figure showing the relative abundance of tax by BMI group.**

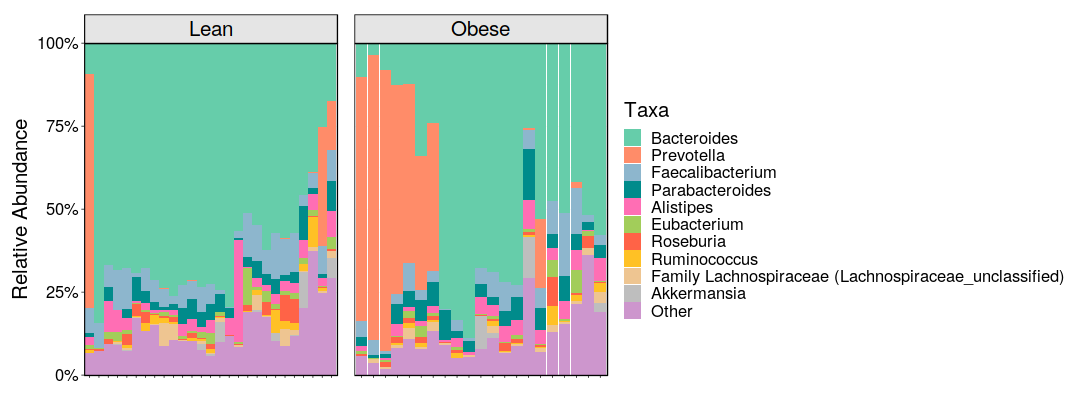
